# Development of a nextflow bioinformatics pipeline for the detection of SARS-CoV-2 co-infection cases from genomic surveillance in the Philippines

**DOI:** 10.1101/2024.07.03.24309627

**Authors:** Adeliza Mae L. Realingo, Francisco Gerardo M. Polotan, Miguel Francisco B. Abulencia, Roslind Anne R. Pantoni, Jessel Babe G. Capin, Gerald Ivan Sotelo, Maria Carmen A. Corpuz, Neil Tristan M. Yabut, Saul M. Rojas, Ma. Angelica Tujan, Karen Iana Tomas, Ardiane Ysabelle Dolor, Czarina Christelle Alyannah Celis, Stephen Paul Ortia, Ezekiel Melo, Chelsea Mae Reyes, Elijah Miguel P. Flores, Anne Pauline A. Alpino, Aldwin Kim A. Penales, Kathlene Mae C. Medina, Joanna Ina Manalo, Timothy John R. Dizon, Katie Hampson, Sandeep Kasaragod, Joseph Hughes, Kirstyn Brunker

**Author notes:** Corresponding author contact details: Adeliza Mae L. Realingo. These authors contributed equally to this work (Adeliza Mae L. Realingo, Francisco Gerardo M. Polotan, Joseph Hughes, Kirstyn Brunker).

## Abstract

Co-infection with multiple SARS-CoV-2 variants, though rare, may have clinical and public health implications, including facilitating variant recombination. Early detection of co-infections is therefore crucial. In this study, we report two probable cases of co-infection identified during routine genomic surveillance. Initially suspected as cross-contamination due to the presence of private mutations and nucleotide mixtures flagged by Nextclade and bammix, the samples were re-extracted and resequenced after workspace decontamination, yet the anomalies persisted. To investigate further, we developed a bioinformatics pipeline (Katmon) incorporating various tools such as Freyja, with lineage abundance results that illustrated presence of multiple variants, and VirStrain, which confirmed inconsistent lineage assignments. We also visualised the alternative allele fractions for each lineage-defining mutation and amplicon, showing evidence of two variants — Delta and Omicron — co-existing within a single amplicon. Amplicon sorting effectively separated reads corresponding to the two variants, and the resulting consensus sequences aligned with their respective lineage assignments. These findings suggest that the first sample, PH-RITM-1395, involved a Delta-Omicron co-infection, while the second sample, PH-RITM-4146, contains both a co-infection and a recombinant variant. To further support the second sample’s recombinant nature, we employed sc2rf, which identified Delta-Omicron breakpoints. Retrospective analysis of 1,078 samples from July 2021 to July 2022, encompassing the period of co-circulation of different variants in the Philippines, flagged four additional co-infection cases, including Delta-Omicron and Beta-Omicron suggesting a lower bound co-infection rate of 0.27%, and 0.19%, respectively. Our findings underscore the critical importance of real-time genomic surveillance and advanced bioinformatics pipelines in detecting SARS-CoV-2 co-infections and variant recombination.

## INTRODUCTION

Since the detection of the first case of COVID-19 in March 2020, the Philippines has experienced distinct epidemic waves marked by surges in COVID-19-related hospitalizations and deaths. The emergence of new variants has underpinned such epidemiological patterns globally, emphasising the critical importance of genomic surveillance. The Philippines has emerged as a leading contributor to SARS-CoV-2 sequencing in Southeast Asia, demonstrating the country’s commitment to strengthening its capacity for genomic surveillance of infectious diseases. This effort has contributed to an enhanced understanding of transmission patterns and the association of distinct SARS-CoV-2 lineages with different phases of the epidemic in the Philippines (Tablizo et al., 2020; Li et al., 2022, Tablizo et al., 2022). Several waves of SARS-CoV-2 variants of concern (VOC), associated with increased transmissibility and immune escape, have been identified during the epidemic. The Delta variant was first detected in May 2021 causing a surge in cases and prompted lockdowns and community quarantine throughout the country. Then, in November 2021, the Omicron variant was initially detected and soon dominated, displacing Delta. Co-circulation of lineages may have allowed recombination events, producing the recombinant lineages, such as the Omicron XBB and sub-variants, FLiRT variants, and XEC, that have been circulating more recently (Tamura et al., 2023; Pipek et al., 2024; Aden & Zaheer, 2024; World Health Organization, 2024).

Co-infection occurs when an individual is simultaneously infected with two distinct variants of the virus, typically during periods when the two variants of concern are co-circulating. In co-infection cases, the presence of multiple variants in a subset of host cells is a potential environment for recombination to occur (Bolze et al., 2022). Epidemiological evidence from Andalusian genomic surveillance (January to May 2022) indicates a correlation between an increase in co-infection cases and a rise in circulating recombinant variants, with the unexpectedly high number of co-infections contributing to the emergence of new Delta-Omicron and Omicron-Omicron recombinants (Perez-Florido et al., 2023). A study by Jackson et al. (2021) described multiple independent recombination events involving UK lineages, emphasising the importance of co-infections during periods of high viral prevalence, which can drive the emergence of new recombinant lineages. Recombination can lead to the emergence of a virus with a novel phenotype by combining genetic material from two distinct variants. This process can result in changes to key viral traits, such as increased transmissibility, immune evasion, or altered disease severity. If the newly formed recombinant virus gains an advantage, it may spread more efficiently in the population, potentially leading to further outbreaks (Carabelli et al., 2023; Turakhia et al., 2022).

A recent study in the Philippines investigating the emergence of the recombinant lineage XBC identified one sample, collected in March 2022, with an allele fraction pattern and within-host mutations indicative of an active co-infection (Pangilinan et al., 2023). Since co-infection is a prerequisite for recombination, this suggests that co-infection cases were already present in the Philippines by early 2022. Genomic surveillance data from the country have also identified XBC sequences from as early as January 2022, a period when Delta was being replaced by Omicron. Phylogenomic methods by Turakhia et al. (2022) identified 589 recombination events in 1.6 million samples publicly available in May 2021. While this represents a relatively low rate on a per-sample basis, it is significant in the context of viral evolution, particularly during periods of high viral prevalence. This emphasises the importance of timely detection and analysis of recombinant lineages to pinpoint their emergence and understand their potential for enhanced epidemiological or phenotypic properties (Turakhia et al., 2022).

Co-infections are rarely reported, but such cases can have important clinical and epidemiological significance, potentially contributing to increased disease severity,and viral immune escape (Bolze et al., 2022; Shiraz & Tripathi, 2022). A study by Rockett et al. (2022), investigated two immunocompromised individuals who experienced co-infection of Delta and Omicron variants. In one case, the patient was initially infected with Omicron and later superinfected with Delta before being admitted to the hospital. Another case study by Samoilov et al. (2021), reported an elderly patient who was hospitalized with worsening symptoms that ultimately led to death. Genomic analysis revealed that the patient was infected with two distinct SARS-CoV-2 strains from different phylogenetic clades, GH and GR. Notably, the relative abundances of these lineages differed between the first and last swabs, taken just eight days apart. The detection of co-infection cases are likely underestimated (Combes et al., 2022), and it is therefore crucial to detect these cases during genomic surveillance as early as possible for clinical management, treatment strategies, and implementing appropriate public health measures.

To accurately detect co-infection cases, it is essential to consider several factors during the sequencing process. Primer amplification bias of some genomic regions of specific variants, contamination during the sequencing process (which might be the actual cause of nucleotide mixtures in the sample), and insufficient bioinformatics methods to systematically detect co-infections can all influence the ability to distinguish true co-infection cases from within-sample noise (Bal et al., 2022). The sample needs to be sequenced at sufficiently high depth to identify and characterise co-infection of different VOCs (Bolze et al., 2022). As variants evolve, both sequencing procedures and bioinformatics tools need critical quality control measures to determine the presence of any co-infection, recombination, and contamination in the sequence data. Currently, there are limited tools for identifying and describing SARS-CoV-2 co-infections of known variants. In Australia, Rockett et al. (2022) were able to flag heterozygous calls for sampled sequences from epidemiologically unrelated patients using their bioinformatic quality control system, similar to the findings that prompted further investigation in our study. In another study in Costa Rica, Molina-Mora et al. (2022), developed a metagenomic pipeline to identify co-infections of distinct SARS-CoV-2 variants as part of their genomic surveillance efforts. A combination of different approaches was required to detect these co-infections, which may vary from case to case.

Co-infections may have been occurring in the Philippines as early as March 2022 (Pangilinan et al., 2023). Here, we investigate two samples, collected in May 2022 and April 2023, flagged as possible Delta and Omicron co-infections during routine genomic surveillance efforts at the Research Institute for Tropical Medicine (RITM). The samples prompted the development of a robust automated pipeline, integrating various independent tools, to diagnose co-infection cases and we retrospectively quantified their prevalence in a large cohort of samples sequenced during the period where Delta and Omicron co-circulated in the Philippines.

## RESULTS

### Initial Discrepancies in Lineage Assignment

Two samples out of 3,254 sequences, as of September 2024, processed by RITM for routine SARS-CoV-2 genomic surveillance were flagged for additional quality control checks due to inconsistencies in their lineage assignments and multiple private mutations (Table 1, Supplementary Figure 1).

**Table 1.** Lineage assignment using Nextclade, Pangolin, and Scorpio.

| Sample |  | Coverage | Nextclade | Pangolin | Scorpio | note |
| --- | --- | --- | --- | --- | --- | --- |
| **PH-RITM-1**<br>**395** | First Sequencing | 99.6% | B.1.617.2<br>(Delta) | B.1.1.529<br>(Omicron) | Probable<br>Omicron<br>(Unassigned) | USHER placements:<br>B.1.1.529(1/1);<br>scorpio found<br>insufficient support<br>to assign a specific<br>lineage |
|  | Replicate | 91.9% | B.1.617.2<br>(Delta) | B.1.1.529<br>(Omicron) | Delta<br>(B.1.617.2-like) | USHER placements:<br>B.1.1.529(1/1);<br>scorpio lineage<br>B.1.617.2 conflicts<br>with inference<br>lineage B.1.1.529 |
| **PH-RITM-4**<br>**146** | First Sequencing | 94.3% | B.1.617.2<br>(Delta) | AY.1<br>(Delta) | Delta<br>(B.1.617.2-like)<br>+K417N | USHER placements:<br>AY.1(2/4),<br>AY.122(1/4),<br>B.1.617.2(1/4) |
|  | Replicate | 99.5% | B.1.617.2<br>(Delta) | AY.1<br>(Delta) | Delta<br>(B.1.617.2-like)<br>+K417N | USHER placements:<br>AY.1(2/4),<br>AY.122(1/4),<br>B.1.617.2(1/4) |

For sample PH-RITM-1395, the consensus sequence had a coverage of 99.57% and was initially identified as B.1.1.529 (Omicron) by the Pangolin tool (O’ Toole et al., 2021), while Scorpio (O’ Toole et al., 2021) determined the sample to be a probable Omicron. This was discordant with Nextclade’s (Hadfield et al., 2018) lineage assignment of B.1.617.2 (Delta). UShER (Ultrafast Sample placement on Existing tRees) (Turakhia et al., 2022) placement noted that this sequence has insufficient support to be assigned to a specific lineage. Meanwhile, the sample PH-RITM-4146 had an initial coverage of 94.3% and was assigned by Pangolin as AY.1 (Delta) and by Nextclade as B.1.617.2 (Delta). The Scorpio call also determined the sample as Delta B.1.617.2-like with an additional K417N mutation, characteristic of a Delta plus variant (Table 1).

Notably, Nextclade documentation notes that sequences with an unusually high number of private mutations are often flagged as potentially erroneous, possibly due to contamination, co-infection, or recombination events (Hadfield et al., 2018).

### Contamination Ruling and Re-Sequencing Confirmation

Despite the flagged lineage assignment inconsistencies, the negative controls from the initial sequencing showed no reads, suggesting that cross-contamination during sample preparation was unlikely. Although collecting multiple samples from the same patient would have been ideal to rule out contamination, this was not possible in our case. Instead, we re-extracted and re-sequenced the samples to confirm the results and eliminate contamination as a factor. Upon doing so, we confirmed that the flagged private mutations persisted, thus ruling out the possibility of cross-contamination.

Further checks for cross-contamination in the wet lab protocol include thorough investigation of the negative extraction control (NEC). The NEC is checked for Qubit concentrations having the acceptable value to be <1 ng/ul. When the Qubit concentration of the NEC exceeds the acceptable value, it is subjected to qPCR to check for amplification of the E and N genes. If the Ct values of the NEC are <40, the entire batch of samples is re-extracted. In the case of PH-RITM-1395, the NEC had a Qubit concentration of 0.4 ng/ul while the NEC of PH-RITM-4146 had a Qubit concentration of 0.120 ng/ul. Therefore, the samples had NECs passing the quality control and cross-contamination was ruled out.

Moreover, as part of the acceptance criteria for sequencing, the Ct value of the specimens were recorded. For the first sample, the Ct values were 19.4 for N gene and 17.2 for E gene. For the second sample, the Ct values were 30.82 for N gene and 19.58 for ORF. These Ct values were low (<40), implying high source viral load that would be less susceptible to contamination, indicating suitability for whole-genome sequencing (WGS).

The results of the replicate consensus sequence for sample PH-RITM-1395 had a coverage of 91.88%, and its lineage assignment remained inconsistent between Pangolin and Nextclade – which showed B.1.1.529 (Omicron) and B.1.617.2 (Delta), respectively. UShER placements also highlighted a conflict in lineage assignment of B.1.617.2 and B.1.1.529.

On the other hand, sample PH-RITM-4146 retained the lineage assignment for Pangolin and Nextclade as AY.1 (Delta) and B.1.617.2 (Delta), respectively. UShER placements indicated the sample to be either AY.1, AY.122, or B.1.617.2.

We then used bammix (Ruis, 2022) which flagged multiple sites with nucleotide base mixtures exceeding our set default 20% minor allele threshold in both samples, as shown in Figure 1. Multiple sites containing heterogeneous bases crossing the threshold in both replicates of each sample is also apparent. To validate these findings, we manually inspected the aligned sequence reads at each position using Tablet software (Milne et al., 2013) and observed the presence of a mixture of nucleotides on the flagged positions. It was apparent that the nucleotide mixtures persisted in the sequencing replicates for both samples. Since we ruled out cross-contamination as the source of lineage assignment discrepancies, we conducted a more thorough investigation for possible co-infection of the two VOCs Delta and Omicron using additional bioinformatics tools.

**Figure 1.**
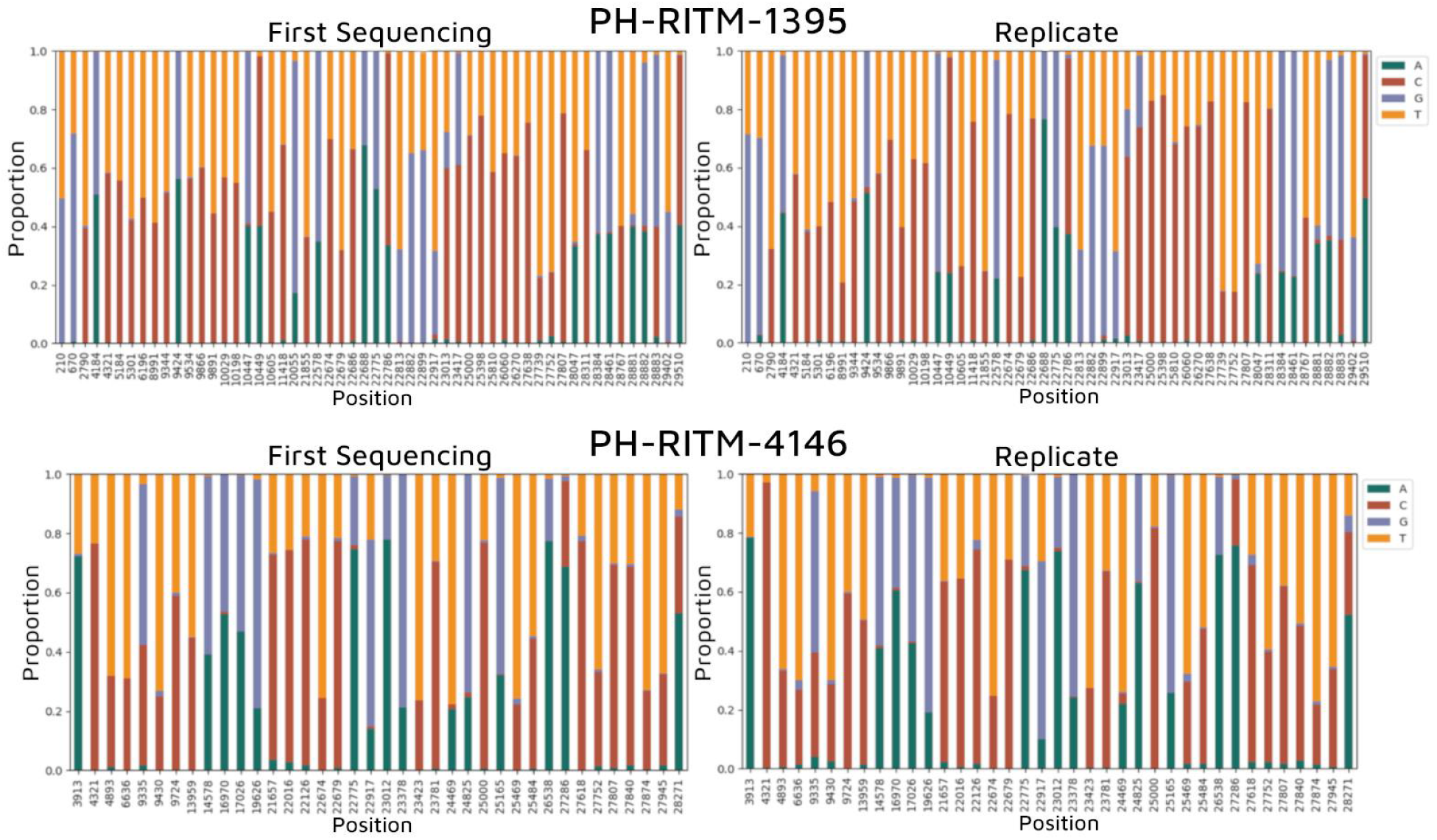
Positions flagged by bammix for nucleotide mixtures for each sample. All sites with single nucleotide variant (SNV) or single N’s, or ambiguous base call, in the consensus assembly of samples of PH-RITM-1395 and PH-RITM-4146.

### Confirmation of Co-Infection

Observation of the nucleotide positions flagged by bammix and examination of overlapping SNVs and mutations support our hypothesis that the sample was from an individual co-infected with Delta and Omicron. To confirm co-infection, we further analysed the sample using VirStrain (Liao, Cai, & Sun, 2022), Freyja (Karthikeyan et al., 2022), and investigated the allele fraction of sequencing reads that mapped to mutations unique to each variant of concern.

#### i) Virstrain

Both samples were run against a custom VirStrain database created from the downloadable multi-fasta file of Variants of Interest (VOI) representative sequences from GISAID (Global Initiative on Sharing All Influenza Data) and the ENA (European Nucleotide Archive) COVID-19 Data Portal. VirStrain identifies the closest relative lineage in the database and can detect two strains with the same clades while providing accurate abundance predictions (Liao, Cai, & Sun, 2022). For PH-RITM-1395, results from VirStrain were consistent with the lineage assignment from both Pangolin and Nextclade: B.1.617.2 (Delta) as the most probable strain, with BA.2 (Omicron) as the other possible strain (Supplementary Files 1 and 2). Meanwhile, for PH-RITM-4146, the most probable strains were AY.4 (Delta) and BA.3 (Omicron), discordant with the results from the two lineage assignment tools (Supplementary Files 3 and 4).

#### ii) Freyja

We then used Freyja, a tool designed to estimate the relative abundances of SARS-CoV-2 lineages in mixed samples from BAM files aligned to the Hu-1 reference. This tool uses lineage-specific mutational barcodes derived from the UShER global phylogenetic tree to address the constrained deconvolution problem, ensuring non-negative values and that the lineage proportions sum to one (Karthikeyan et al., 2022).

For the initial sequence of PH-RITM-1395, abundances of Delta and Omicron are almost the same for the first and replicate sequences, and other variants were reported but at low levels. The estimated lineage abundance of Delta and Omicron was 49.81% and 48.09%, respectively. The replicate sequence had a slight increase in Delta abundance, 56.83% as compared to Omicron with 42.55%. Other variants were detected by Freyja but at low levels (Figure 2; Supplementary File 5).

**Figure 2.**
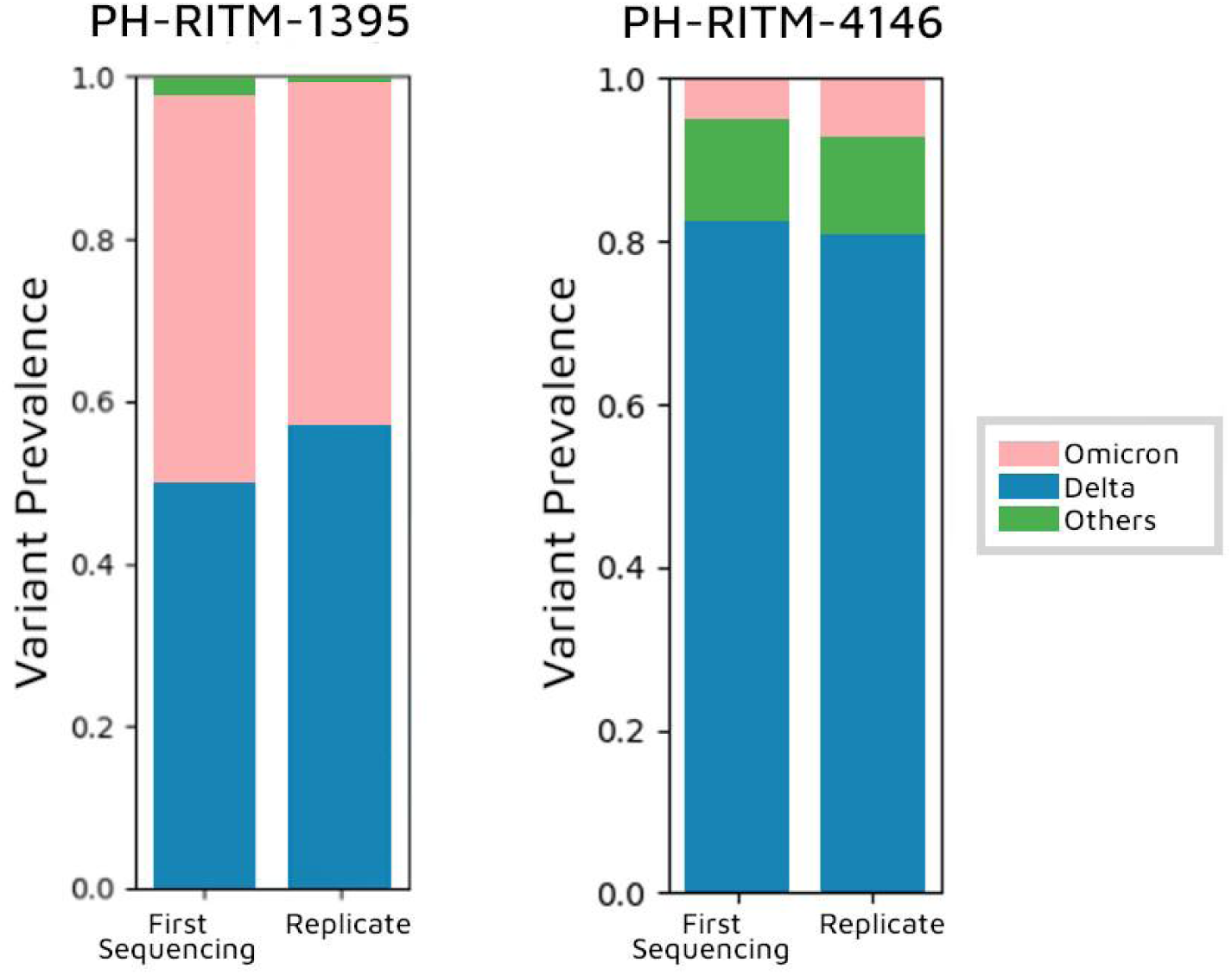
Freyja variant prevalence results. Variant frequency in the samples PH-RITM-1395 and PH-RITM-4146 for the first and replicated sequence run plotted by Freyja tool.

For sample PH-RITM-4146, the majority of the sequences were classified as Delta, with a smaller proportion identified as Omicron. Interestingly, Freyja detected a relatively higher prevalence of “other variants,” primarily composed of recombinant lineages, accounting for 12.24% of the total abundance. In the replicate sequence, the abundance of these “other variants” slightly decreased to 11.87%.These ‘other variants’ include the recombinant lineages XAC and XZ, derived from Omicron sublineages BA.2 and BA.1; XD a recombinant of B.1.617.2 (Delta) and BA.1 (Omicron); and XAY, a recombinant of AY.45 and BA.4/BA.5 (Figure 2, Supplementary File 6). This suggests the sample contains both Delta and Omicron variants as well as recombinant variants.

#### iii) Comparative Allele Fraction Analysis

From cov-spectrum.org, we listed the established mutations that are shared and distinct to B.1.617.2 (Delta) and B.1.1.529 (Omicron). Delta has 74 lineage-defining mutations, while Omicron has 51. Additionally, four amino acid changes are shared between the two namely, ORF1b:P314L, S:S477N, S:T478K, and S:D614G. Similar to the co-infection analysis described by Bolze et al. (2022), we utilised custom python scripts illustrating the alternative allele fraction of the mutations across the SARS-CoV-2 genome of both samples (Figure 3). Only high-quality reads (Q 20) were used to create the allele fraction plots. Mutations shared between the two variants have an allele fraction equal to one, as expected since the allele is found in all of the reads in that position.

**Figure 3.**
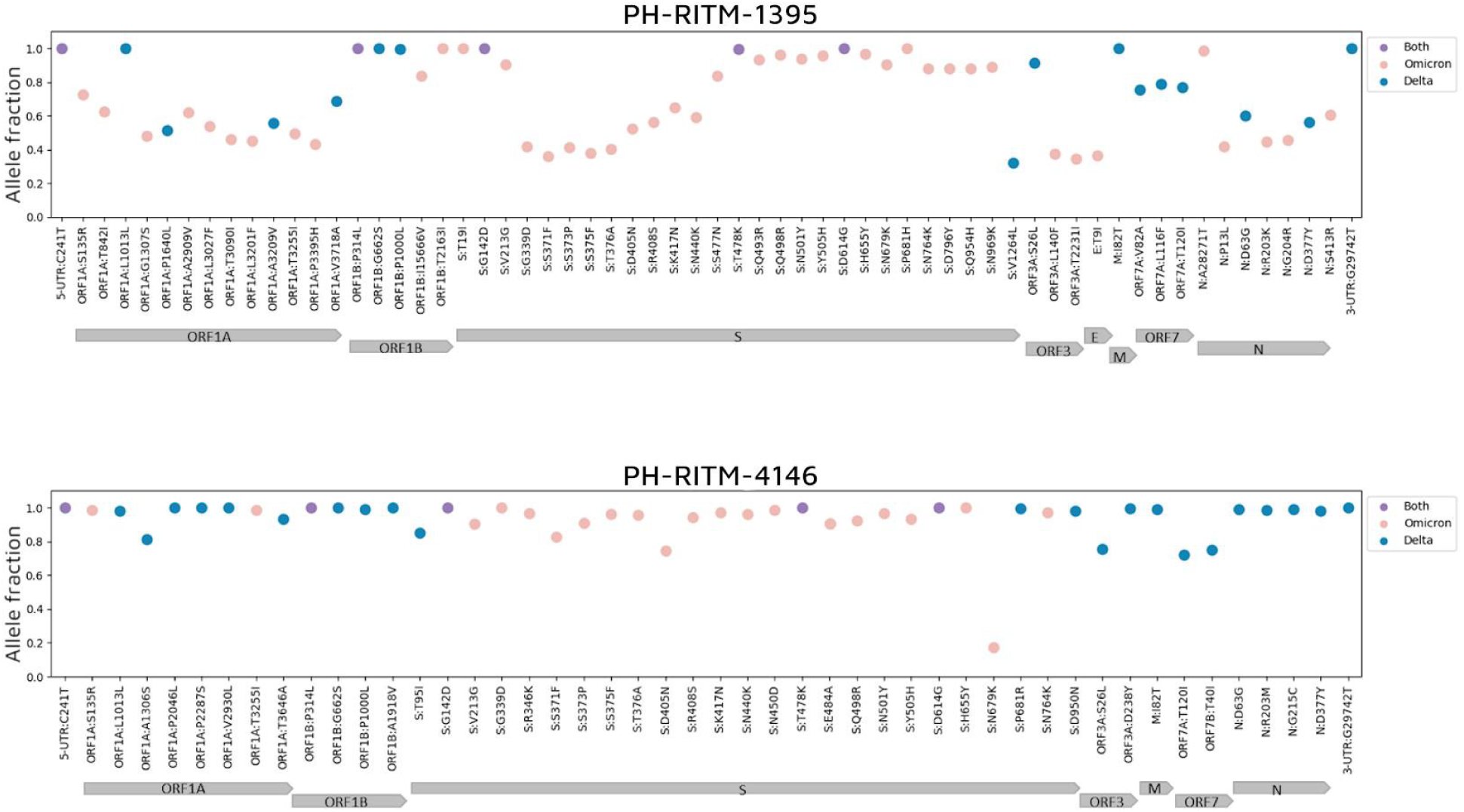
Alternative allele fraction of PH-RITM-1395 and PH-RITM-4146 for each lineage-defining mutation across the SARS-Cov-2 genome. Delta, Omicron, and shared mutations are represented by blue, pink, and purple, respectively.

As shown in figure 3, the presence of both Delta and Omicron defining mutations in PH-RITM-1395 supports the hypothesis that this sample represents a co-infection case, as the mixed alleles exhibit fractions below one, which is a pattern consistent with co-infection (Bolze et al., 2022). In contrast, sample PH-RITM-4146 exhibits Delta and Omicron lineage-defining mutations in distinct genomic regions, with allele fractions close to one and a sudden drop in allele fraction at the Spike mutation N679K, a pattern characteristic of recombinants also described by Bolze et al. (2022).

We then determined the AAF for each amplicon. PH-RITM-1395 shows a mixture of the two VOCs for almost all amplicons, supporting the pattern of co-infection. In contrast, PH-RITM-4146 displayed minimal mutation mixtures within amplicons, with distinct genomic regions showing amplicons corresponding exclusively to either Delta or Omicron. This pattern strongly supports the sample as a recombinant variant (Figure 4).

**Figure 4.**
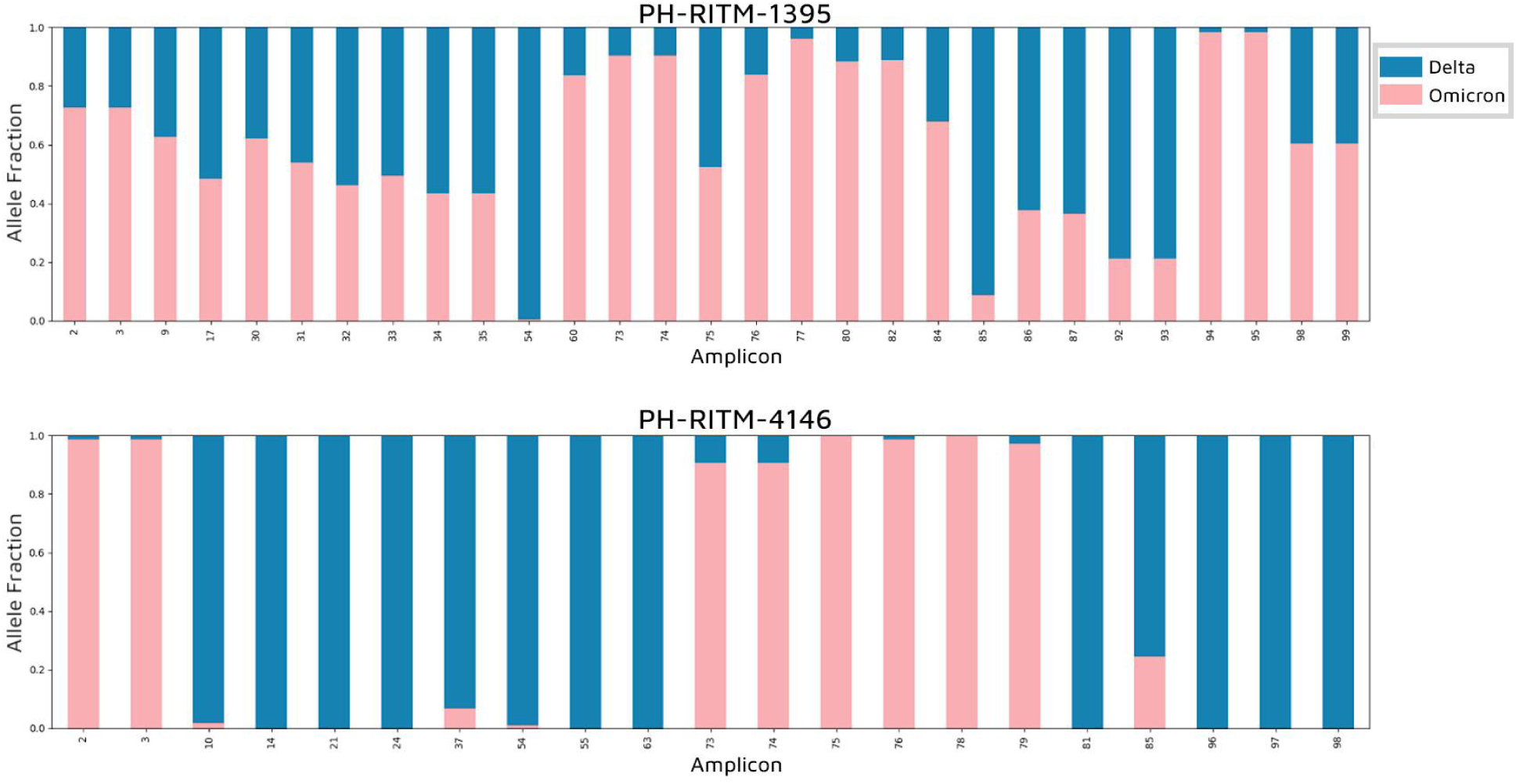
Alternative allele fraction per amplicon. The fraction of reads per amplicon having Delta (blue) and Omicron (pink) lineage defining mutations were plotted.

Both the AAF plots per lineage-defining mutation and per amplicon support the hypothesis that PH-RITM-1395 contains a mixture of reads from both Delta and Omicron variants within a single amplicon, suggesting co-infection. In contrast, the AAF plots for PH-RITM-4146 show a pattern more indicative of recombination, with distinct regions of amplicons associated with either Delta or Omicron, possibly pointing to breakpoints within the sample.

The *t*-test analysis results showed that for the replicate sequence of PH-RITM-1395, no significant differences were observed in the proportions of Delta and Omicron for both the first (*p* = 0.73) and replicate (*p* = 0.69) sequences. Therefore, the variant prevalence for Delta (blue) and Omicron (pink) remained almost similar after the first and replicate sequencing. These findings rule out contamination as the source of mixed variant mutations, further supporting PH-RITM-1395 as a co-infection case.

#### iv) Amplicon Sorting

To further demonstrate the presence of mixed reads belonging to Omicron and Delta within a single sample, we sorted the reads associated with each variant. Utilising the samjdk tool in jvarkit (Lindenbaum & Redon, 2017) for amplicon sorting, we effectively separated reads from the BAM file by identifying the lineage-defining mutations present in each read. Results from running custom python scripts for amplicon sorting in Table 2 showed the assigned lineages for sample PH-RITM-1395 as B.1.617.2 (Delta) and BA.2 (Omicron) for the Pangolin tool, while Nextclade assigned them as B.1.617.2 (Delta) and BA.2.3 (Omicron). These results are concordant with VirStrain. On the other hand, the sorted sequences of sample PH-RITM-4146 were assigned as AY.1 (Delta) and B.1.1.529 (Omicron) by Pangolin, while Nextclade classified them as B.1.617.2 (Delta) and B.1 (Omicron). The lineage assignment for the Delta-specific reads in PH-RITM-4146 aligns with the initial results from both Pangolin (AY.1) and Nextclade (B.1.617.2). These findings further support the presence of reads originating from both Delta and Omicron within the sample, also underscoring the value of employing multiple tools to achieve a more comprehensive and reliable lineage assignment in samples suspected for co-infection.

**Table 2.** Lineage assignment after amplicon sorting.

| Sample | Nextclade | Pangolin | Scorpio | note |
| --- | --- | --- | --- | --- |
| PH-RITM-1395 | B.1.617.2 | B.1.617.2 | Delta<br>(B.1.617.2-like) | UshER placements: B.1.617.2(1/1) |
|  | BA.2 | BA.2.3 | Probable<br>Omicron<br>(Unassigned) | UShER placements: BA.2(1/1) |
| PH-RITM-4146 | AY.1 | B.1.617.2 | Delta<br>(B.1.617.2-like)<br>+K417N | UShER placements: AY.1(2/4),<br>AY.122(1/4), B.1.617.2(1/4) |
|  | B.1.617.2 | B.1 | Probable<br>Omicron<br>(Unassigned) | UShER placements:<br>B.1.1.529(2/2); scorpio found<br>insufficient support to assign a<br>specific lineage |

### Confirmation of Recombinant Variant in PH-RITM-4146

Due to the allele fraction pattern for PH-RITM-4146, we considered the sample to be a recombinant and subjected it to the sc2rf tool (Schimmel et al., 2022). Sc2rf is a command-line program specifically designed for detecting potential SARS-CoV-2 recombinants by identifying regions in the genome where abrupt changes in lineage-defining mutations occur.

The tool detected four breakpoints in PH-RITM-4146, suggesting a recombinant between Delta (21J) and Omicron (BA.2 or 21L) (Supplementary Figure 2). The consistent detection of breakpoints across multiple runs of the tool, as well as the alignment of these breakpoints with lineage-specific mutations, supports the reliability of the recombination signal. Additionally, the location of the breakpoints correspond to genomic regions where recombination events have been previously reported in SARS-CoV-2, further supporting the validity of the identification.

To strengthen the evidence for recombination, we also verified the presence of lineage-defining mutations on either side of the breakpoints, confirming the alternating patterns consistent with Delta and Omicron lineages. This pattern is characteristic of a recombinant virus, distinguishing it from a co-infection, which would exhibit overlapping or blended allele fractions rather than clear transitions between distinct variant-defining mutations.

The sc2rf results further support the Freyja analysis, which detected “other variants” with an average abundance of 12%, indicating the presence of recombinant lineages in the sample. Specifically, Freyja identified signals consistent with recombination between Delta and Omicron, corresponding to the detection of XD, a known Delta-Omicron recombinant. The consistent identification of recombinant signals by both Freyja and sc2rf makes it unlikely that the “other variants” detected by Freyja are due to misclassification.

### Pipeline development, validation on simulated mixtures and retrospective application to routine surveillance samples

We developed a pipeline compiling the steps used to analyse these two flagged samples (Figure 5). The pipeline is designed as a general tool for analysing sequences generated from routine SARS-CoV-2 genomic surveillance. It integrates community-standard tools to deliver comprehensive results, helping users to assess lineage discrepancies and identify potential co-infections or recombinant cases. The pipeline generates a summary report as an output, which includes the results from lineage assignment, flags for nucleotide mixtures, and the alternative allele fraction plots.

**Figure 5.**
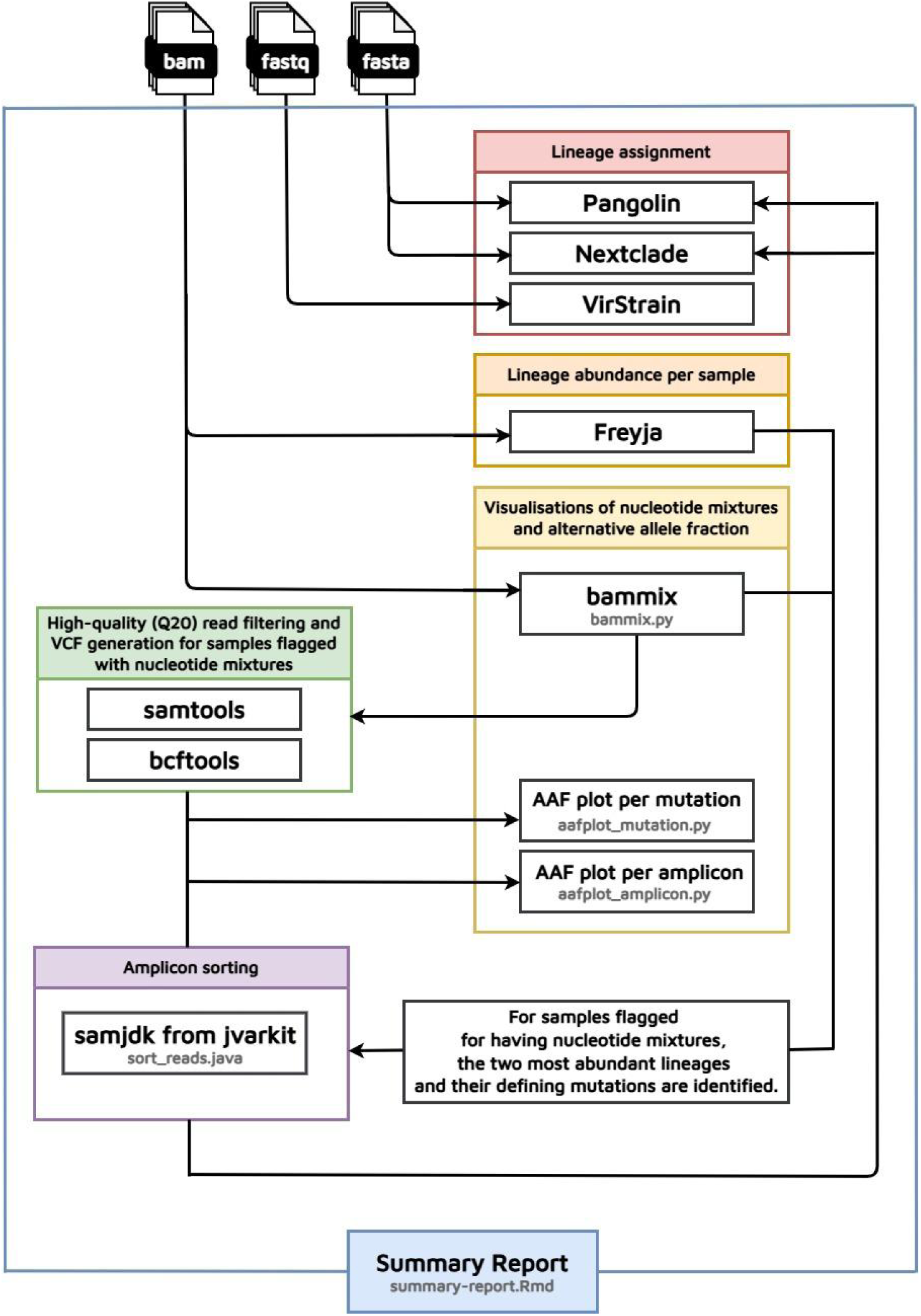
Flow of the Katmon pipeline developed. The pipeline developed to analyse samples that were flagged as probable co-infection used various tools for lineage assignment, lineage abundance, nucleotide mixtures visualisations, and amplicon sorting. Some of the steps used custom python, java, and R scripts. The pipeline outputs all the results per step in sorted folders and creates a summary report.

To validate the pipeline’s applicability in detecting co-infections, we ran Katmon on simulated reads of randomized lineage combinations at varying proportions of major and minor allele (50:50, 80:20, 90:10, 95:5, and 98:2). Our results show that the pipeline can detect co-infections with minor alleles as low as 2% by adjusting the bammix threshold for identifying nucleotide mixtures. Additionally, Freyja’s ability to specify lineage abundances enables the detection of co-infections involving variants from the same clade.

The subsequent application of the Katmon pipeline to archived samples further identified four potential co-infections, as shown in Figure 6, with the earliest case dating back to December 2021. This brings the total number of co-infection cases in the Philippines to five out of 1,078 samples processed between July 2021 and July 2022. Among these, three cases involved co-infection with Delta and Omicron, while two involved Beta and Omicron. Hence, the prevalence of Delta and Omicron co-infections was 0.27%, and Beta and Omicron co-infections had a prevalence of 0.19%.

**Figure 6.**
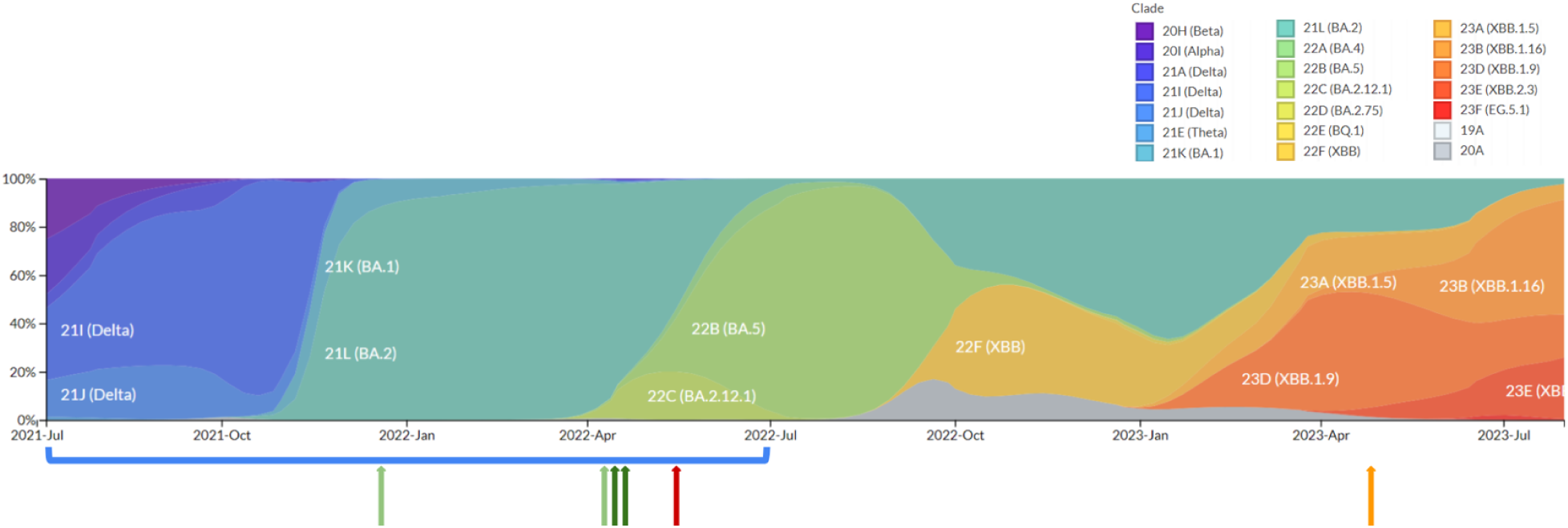
Lineage frequency plot of the Philippine SARS-CoV-2 sequences. The frequency plot illustrates lineage assignment, colored by clade, for all publicly available Philippine samples collected from July 2021 to July 2023. The red and orange arrows, representing PH-RITM-1395 and PH-RITM-4146, respectively, indicate the samples initially investigated for co-infection, and are the basis of the bioinformatics pipeline development. Out of 1,078 archived samples analysed, from the period of July 2021 to July 2022 (blue bracket), the pipeline detected a total of five possible co-infection cases: the initial sample PH-RITM-1395 (red arrow), two cases of Delta and Omicron co-infection (light green arrows), and two cases of Beta and Omicron co-infection (dark green arrows).

Notably, the Delta-Omicron co-infected samples originated from different batches and regions, with the first one collected from the National Capital Region (NCR) in December 2021 and the other one from Zamboanga (Region IX) in April 2022. Both collection dates predate the sample PH-RITM-1395 investigated in detail above, which was also from Zamboanga (Region IX). Meanwhile, the Beta-Omicron co-infected samples were collected within three days of each other in April 2022, from both the same batch and region (NCR).

## DISCUSSION

In this study, we identified two potential cases of co-infection between SARS-CoV-2 variants Delta and Omicron through routine genomic surveillance during the COVID-19 pandemic in the Philippines. The two samples, PH-RITM-1395 and PH-RITM-4146, exhibited discordant lineage assignments and an unusually high number of private mutations, prompting further investigation for possible contamination, co-infection, or recombination. After re-extracting and resequencing the samples to rule out cross-contamination, we developed and implemented a custom bioinformatics pipeline. At the time of genomic surveillance, existing processes for detecting co-infections were limited, so we employed additional bioinformatics tools for a more stringent analysis of nucleotide mixtures within the samples. This thorough investigation enabled us to rule out cross-contamination and distinguish between cases of co-infection, and the presence of two distinct variants and recombinants within a sample. Results from the simulated data demonstrate that the developed pipeline can detect co-infections with minor variants as low as 2% and identify co-infections of lineages belonging to the same clade.

Applying the refined pipeline to archived samples, we uncovered four additional cases, including Delta-Omicron and Beta-Omicron co-infections. Our findings demonstrate that the persistence and co-circulation of different variants did result in co-infections, presenting the opportunity for recombination and the emergence of recombinant lineages.

Cases of co-infection between Omicron and Delta have been found in many countries namely Australia, Argentina, Equatorial Guinea, France, Singapore, Spain, South Africa, and the USA (Rockett et al., 2022; Pisano et al., 2022; Hosch et al., 2021; Combes et al., 2022; Bal et al., 2022; Liu et al., 2021; Perez-Florido et al., 2023; Pipek et al., 2024; Zhou et al., 2022). Most of these however were from cases during the lineage replacement of Delta by Omicron from late 2021 to early 2022, whereas our case was detected near the middle of 2022 in May, a point in time in the Philippines when Omicron had become dominant, and Delta had reportedly only two cases during the said month. It is also possible that the patient had a chronic infection that started earlier (Pedro et al., 2021). Interestingly, the latest Delta variant Philippine sequence available on GISAID was collected from Davao in January 2023 and submitted by the Philippine Genome Center (PGC). A small number of Delta variant sequences (*n* = 50) were still being collected in 2023 from countries in Africa, Asia, Europe, and North America according to sequence data from GISAID (Khare et al., 2021). Unfortunately, at this time, we have been unable to confirm whether these are collection date misannotations during submission.

Sample collection was only performed at one point in time, as this was done prior to the development of our pipeline to automatically flag anomalous reads, so we were unable to observe any possible changes in relative abundance in the two variants over time. In a study by Pedro et al. 2021, co-infection by two SARS-CoV-2 lineages was detected in a patient wherein the frequency of the lineage changed over time, from an absolute 20A lineage at the time of diagnosis, to a 3% 20B lineage frequency after nine days, and then to a 100% 20B lineage frequency after two months. In a similar study done by Samoilov et al. (2021), in which swabs from the same patient, only 8 days apart, were reported to have different lineage abundances of clades GH and GR. This indicates the importance of temporal sampling when it comes to monitoring the evolution of viral populations and might be a point for consideration in improving genomic surveillance, particularly in patients with chronic COVID-19 infection (Samoilov et al., 2021; Pedro et al., 2021; Rockett et al, 2022).

The two cases presented here stood out as anomalies during routine genomic surveillance, triggering an investigation that confirmed co-infection. Genomic surveillance enables the estimation of the prevalence of co-infections and thus the risk of emerging recombinant lineages. Perez-Florido et al. (2023) reported that the unexpectedly high frequency of co-infections could lead to the emergence of new recombinant variants as they were able to define two new recombinant samples. Besides the risks of new recombinant variants, certain specific co-infections may sometimes lead to more severe disease, resulting in death (Samoilov et al., 2021; Bal et al., 2022). If done in a timely manner, it can potentially inform diagnostics, proper patient care, and improve clinical outcomes. To address this need, we developed a bioinformatics pipeline designed to flag and analyse possible co-infection cases. Recent developments have expanded the pipeline’s capabilities to detect any SARS-CoV-2 variants co-infection.

In the Philippines, we found that the prevalence of Delta and Omicron co-infections was 0.27% while the prevalence of Beta and Omicron co-infections was 0.19%. Estimates of the prevalence of co-infections have varied worldwide, potentially due to a multitude of factors including the varying genetic diversity of SARS-CoV-2 lineages circulating at the time of study, differences in transmission between different geographical locations, and methodological differences (Bal et al., 2022; Liu et al., 2021; Tonkin-Hill et al., 2021; Dezordi et al., 2022; Pipek et al., 2024). For example, in a cohort study in Singapore from January to March 2020, a prevalence of 8% was reported among 131 patients (Liu et al., 2021). In the USA, the reported rate of co-infection is 0.3%-0.5% from January to September 2021 (Zhou et al., 2022) and a similar prevalence of 0.35% co-infection was observed in South Africa (Pipek et al., 2024). In France, co-infection with Delta and Omicron (BA.1), and BA.1 and BA.2 Omicron lineages, were estimated to have a lower prevalence of 0.18% and 0.26%, respectively, for the period December 2021 to February 2022 (Bal et al., 2022); These percentages are almost similar to those we have calculated for the Philippines. Another study in France that screened 3237 samples for VOC-specific SNPs observed 0.2% of samples co-infection with Delta and Omicron and even detected a recombination event between the two variants (Combes et al., 2022). In Argentina, there was a 0.1% prevalence of Delta-Omicron co-infection over a year period from December 2021 to January 2022 (Pisano et al., 2022). Meanwhile, the first case of dual infection with other VOCs, namely Beta and Delta, was reported in Equatorial Guinea with an overall prevalence of co-infection in the country at 2.1% (Hosch et al., 2021), highlighting the possibility of co-infection with any combination of variants. Overall, these findings indicate that while co-infections with SARS-CoV-2 variants have been documented across different regions, their prevalence remains relatively low, suggesting that such cases are uncommon despite the concurrent circulation of multiple lineages. However, this is potentially due to the relatively small number of studies, the limited sensitivity of detection methods, under-sampling, or the challenges in identifying mixed infections from sequencing data.

The distinction between co-infection and recombinant variants is evident in the observed alternative allele fraction patterns of the samples that we investigated. Co-infections typically show mixed alleles from multiple lineages throughout the genome, while recombinant variants display lineage-defining mutations in distinct genomic regions. Sample PH-RITM-4146 also exhibited a significant number of flagged private mutations identified by Nextclade; however, the overall analysis supports that the sample is a recombinant rather than a case of co-infection. Sc2rf results suggest that PH-RITM-4146 contains breakpoints pointing to Delta (21J) and Omicron (BA.2 or 21L), thus strongly suggests that it has undergone a recombination event. This conclusion is supported by the predominance of recombinant variants circulating in April 2023, when the sample was collected. Interestingly, the sample involves both Delta and Omicron, despite the fact that no Delta cases had been reported in the Philippines since January 2023. As sequencing high-quality longer reads becomes more cost-effective and widely available, it will be a useful method that enables better discrimination of recombinants. Other studies have reported the characterization of co-infections and recombinant variants using single-molecule real-time sequencing (PacBio technology), demonstrating its potential for resolving complex viral populations with greater accuracy (Trémeaux et al., 2023; Combes et al., 2022). Applying this approach to the sample could help distinguish whether the observed signals represent a true recombinant variant or a mixture of parental and recombinant variants.

The Delta-Omicron co-infected samples were collected from distinct regions and time points: one from NCR in December 2021 and the other from Zamboanga region in April 2022. Notably, the NCR sample was collected earlier than the PGC sample from the same region thus co-infection cases were occurring as early as December 2021 when Omicron cases started to increase while co-circulating with Delta. Additionally, the Zamboanga sample exhibits similarities to PH-RITM-1395, collected in the same region in May 2022. The limited sample collection from other regions may have contributed to the predominance of reported co-infection cases in these two areas.

## CONCLUSIONS AND RECOMMENDATIONS INCLUDING FUTURE DIRECTIONS

We identified and characterised two probable co-infection cases during routine SARS-CoV-2 genomic surveillance in the Philippines. By implementing stringent decontamination protocols, followed by re-extraction and resequencing, we confidently ruled out the possibility of cross-contamination. Subsequently, our custom bioinformatics pipeline then determined that the first sample represented a co-infection between the dominant Omicron variant and a persistent Delta lineage, while the second sample contained both Delta and Omicron variants and recombinant variants, indication co-infection with evidence of recombination.

This is the first report of a confirmed Delta-Omicron co-infection case in the Philippines, detected during a period when the two variants were co-circulating. Meanwhile, despite the absence of circulating Delta variants at the time of sample collection of the second sample, it is possible that the recombinant strain emerged during the co-circulation of Delta and Omicron and persisted through transmission between individuals, remaining undetected in the community even after the Delta variant had largely disappeared. Additionally, a retrospective analysis using our pipeline identified four more probable co-infection cases highlighting the importance of implementing practices to detect and monitor such evolutionary events.

Our initial investigation of the two cases prompted the development of our Katmon pipeline, which streamlines the detection of co-infection cases in routine sequencing efforts. This generates an HTML report summarising lineage assignment, flagged positions with nucleotide mixtures, alternative allele fractions patterns, and results of the amplicon sorted reads – all derived from a combination of community-standard tools and custom scripts. This report will serve as a valuable resource for researchers and public health officials in monitoring and analysing co-infection cases.

Future work should focus on investigating both recent and archived samples, particularly from under-sampled regions of the country, to determine whether cryptic local transmission allowed the Delta lineage to persist despite being replaced by Omicron. Retrospective analyses could also uncover previously unrecognised co-infections. Prospective detection of co-infections through routine integration of the Katmon pipeline could enhance real-time genomic surveillance, enabling the early identification of precursors to recombination, which may drive the emergence of new variants and affect patient outcomes or outbreak trajectories. This would elevate genomic surveillance into a more actionable and informative public health tool.

## MATERIALS AND METHODS

### Sample collection, RNA inactivation and extraction

Samples obtained from naso- and oro-pharyngeal swabs of COVID-19 positive patients were endorsed by the Epidemiology Bureau (EB) to the Research Institute for Tropical Medicine (RITM) for sequencing as part of the routine SARS-CoV-2 genomic biosurveillance in the Philippines. RNA was inactivated and extracted using MagMax^TM^ Viral/Pathogen Nucleic Acid Isolation Kit (A42352, ThermoFisher) in the Kingfisher flex purification system. This protocol used 200 uL of clinical specimen and a final elution volume of 80 uL.

As part of the sequencing protocol, the identifiers used for each sequence were assigned in the laboratory during processing and cannot be directly traced to patients from whom the samples were collected. Among all the samples, we further investigated two sequences, PH-RITM-1395 and PH-RITM-4146, as possible co-infection cases because of their inconsistent lineage assignment and unusual number of private mutations flagged by Nextclade (v2.14.1). The first sample was collected in May 2022 in Zamboanga City, Zamboanga del Sur while the second sample was collected in April 2023 in Daraga, Albay. Since the specimens were endorsed by EB, no serology or RT-qPCR testing for other respiratory pathogens were performed.

### Whole-genome sequencing using Oxford Nanopore Technology (ONT)

Sequencing libraries were prepared using the ARTIC LoCost sequencing protocol^Ⓡ^ (Quick, 2020). The cDNA was prepared by reverse transcription using LunaScript RT^Ⓡ^ SuperMix Kit (E3010, NEB) and ARTIC primers v4+4.1 were used with Q5 Hot Start High-Fidelity 2X Master Mix (M0494, NEB) to amplify the whole genome in a multiplex PCR reaction, followed by purification of the DNA using 0.9X AMPure XP beads (A63881, Beckman Coulter). Sample concentration was checked using a Qubit 4 fluorometer, with acceptable concentrations of >10 ng/ul. To prepare for ONT whole-genome sequencing the amplicons were library prepared and barcoded using Ultra II reagents (E7546 and E7595, NEB) and ONT Native Barcoding Kit (EXP-NBD196), cleaned up with 0.4X AMPure XP beads, and the adapters ligated using the NEBNext Quick Ligation Module (E6056, NEB). The final libraries were prepared using the ONT Ligation Sequencing Kit (SQK-LSK109) and loaded into an R9.4.1 flow cell (FLO-MIN106D, ONT). Sample PH-RITM-1395 was sequenced on a GridION with high accuracy basecalling for 72 hours, while sample PH-RITM-4146 on a MinION Mk1B device with fast basecalling for 48 hours.

### Detection of SARS-CoV-2 variant of concern and pango-lineage

Following the RITM routine process for SARS-CoV-2 genomic surveillance, we used the ARTIC bioinformatics standard protocol for SARS-CoV-2 implemented in the ncov2019-artic-nf nextflow workflow (Artic Network, 2020) to generate a consensus sequence and assign a pango-lineage to the samples. Basecalling of raw ONT fast5 data was performed on the GridION machine using guppy software (v5.1.13). Raw fast5 and fastq files were transferred to the RITM high-performance computing (HPC) server. The workflow performs demultiplexing of samples by barcodes, read filtering by length of 400-700 bp, and mapping of basecalled reads to raw fast5 signal data. Fastq reads are then aligned to the SARS-CoV-2 reference genome using minimap2. Primers are trimmed from aligned reads, and sequences of aligned reads falling outside primer boundaries are soft-clipped. Variants are called using nanopolish for each primer pool separately. Variant calls are merged into a single VCF file, and then filtered with the criteria of >20x depth and a minimum of 50% supporting reads from each strand. Regions of the genome covered with <20x depth are masked, and variant calls are introduced to the regions with >20x depth to produce the consensus sequence. Pangolin tool, including Scorpio (v0.3.17), with default UShER-1.2 as lineage assignment engine was used to assign the consensus sequence to a SARS-CoV-2 lineage and Nextclade (v2.14.1) was used for quality control. As part of genomic surveillance, all the sequences having greater than 70% genome coverage, including the samples investigated in this study, were shared publicly to the Global Initiative for Sharing All Influenza Data (GISAID).

### Detection of Omicron and Delta co-infection

A close examination of the sequence with anomalous quality control metrics was performed, including inspection of the quality of the raw reads, to the consensus sequence, variant calls, and read-level alignment. FastQC (v0.11.9) was used to evaluate the quality of basecalled Fastq reads. The Nextclade (v2.14.1) web server was used to evaluate the quality of the consensus sequence, including metrics such as percent coverage, depth, unexpected private mutations, frameshifts, and stop codons. The bammix tool was used to visualise and identify all the genome positions with heterogeneous nucleotide bases from aligned reads, i.e. not a single dominant nucleotide base but a mixture. The covSPECTRUM and CoVariants web resources were used to identify the list of single nucleotide variants (SNVs) that were distinct between the mixed lineages suspected to be present in the same sample. Multiple SNVs that were located near enough each other and could be spanned by reads that are 400-700 bp long were inspected in the alignment BAM file using Tablet software (v1.21.02.08) (Milne et al., 2013). Individual reads in Tablet containing the expected SNVs of one of the two lineages but not the other were identified as supporting evidence that multiple lineages, and not a single recombinant lineage, was present in the sample. We used VirStrain (v1.17) to confirm lineage inconsistencies from the FASTQ files and Freyja (v1.5.1) to estimate lineage abundance from the BAM files.

We then looked at the alternative allele fraction (AAF), using custom python scripts adapted from Bolze et al. (2022), to determine the number of reads supporting each VOC. AAF is the ratio between the number of reads supporting an alternative allele or mutation and the total number of reads covering the corresponding position. Also using custom python scripts, we illustrated the average AAF of Delta and Omicron per amplicon by first filtering high quality reads from the BAM files and determining the proportion of lineage-defining mutations. The range of each amplicon was defined by a BED file from the ARTIC Network’s version 4.1 primer scheme, available at https://github.com/artic-network/artic-ncov2019. Lastly, using the tool samjdk from jvarkit (Lindenbaum & Redon, 2017), we separated the Omicron and Delta reads, created independent consensus sequences from the sorted reads, and determined their lineage assignment.

### Pipeline development

Following the steps of the manual investigation of the two samples, we developed the Katmon pipeline (Figure 5). The pipeline is written using Nextflow, allowing multiple processes to run in parallel for efficient analysis (Ewels et al., 2020). It is publicly accessible at https://github.com/lanadelrea/Katmon. As input, the pipeline takes BAM, FASTQ, and FASTA files produced during genomic surveillance. The use of the different file types is primarily driven by the varying input requirements of different tools and enables us to examine the sample from multiple perspectives. The pipeline consists of six key steps: (1) lineage assignment using the tools Pangolin, Nextclade, and VirStrain; (2) lineage abundance estimation using Freyja; (3) visualisation of nucleotide mixtures with bammix (4) plotting of alternative allele fraction using filtered high quality reads (Q20); and (5) amplicon sorting of reads, using the tool samjdk from jvarkit, based from lineage-defining mutations of the two most abundant lineage detected from Freyja. Custom scripts were written for generating the AAF plots and performing amplicon sorting. Finally, the pipeline creates a comprehensive summary report that integrates all tables and plots from these processes.

Having a method that independently utilizes different tools and then assesses their agreement provides a significant advantage because each tool possesses their own limitations. Since Pangolin (O’ Toole et al., 2021) and Nextclade (Hadfield et al., 2018) assign lineages by only using the consensus FASTA, we cannot directly confirm the presence of co-infection in the sample. For initial investigation, we utilized Nextclade’s feature that flags positions with private mutations. According to Nextclade documentation, an unusually high number of private mutations may indicate contamination, co-infection, or recombination. We then use bammix (Ruis, 2022) to determine whether these flagged positions contain nucleotide mixtures; however, it cannot determine the specific variants to which the mutations belong to. Then, to compare lineage assignments based on raw FASTQ file reads, we use VirStrain, a strain-level analysis and haplotype reconstruction tool. Its limitations are that it cannot accurately detect low abundance viral strains from very short reads (depth <10x, read length <100bp) and relies on a custom database that must be updated whenever new variants emerge (Liao et al., 2022). In contrast, Freyja, using the BAM files as input, can resolve lineages even at 5% abundance, but requires greater sequencing depth to obtain a more accurate estimate of the true mutation frequency in the samples (Karthikeyan et al., 2022). The Katmon pipeline integrates these multiple analytical tools, examining data from different perspectives to allow the identification of congruent signals, thereby providing a reliable and conservative estimate of co-infection.

The pipeline was initially designed to report possible Delta and Omicron co-infections. We worked on more recent developments to expand its capability in detecting potential co-infections of any SARS-CoV-2 variants – including co-infections of newer variants, and cases involving two lineages of the same clade. To achieve this, we leveraged the ability of Freyja to identify the two most abundant lineages in a sample and retrieve their lineage-defining mutations. These mutations are then used to plot alternative allele fractions and perform amplicon sorting, which improves on the previous method of only sorting reads specific for Delta and Omicron. Additional improvements include enhancing the sensitivity of bammix in detecting nucleotide mixtures. The default threshold for detecting minor alleles is set at 20%, but we have enabled the detection of minor alleles as low as 2%. This threshold can be adjusted by the user as needed.

### Simulation using *in silico generated* reads

To test the pipeline’s ability to detect low-abundance mixtures and co-infections of lineages belonging to the same clade, we simulated mixed reads of randomized lineages mimicking co-infections using ww_simulations (https://github.com/CFSAN-Biostatistics/ww_simulations). The generated reads were combined in various proportions: 2:98, 5:95, 10:90, 20:80, and 50:50. These simulated reads, along with the scripts used to automate read simulation, are available in a separate repository (https://github.com/lanadelrea/simKatmon/). The Katmon results for the simulated reads can also be found in the latter repository.

## DATA AVAILABILITY

All sequences processed for Philippine SARS-CoV-2 genomic surveillance, including those used in this study, have been uploaded to and are publicly available on the EpiCoV database of the Global Initiative for Sharing All Influenza Data (GISAID). Batches containing the flagged samples analysed in this study are accessible in GISAID under the compiled EPI SET (EPI_SET_240703bc), as detailed in Supplementary File 7, and in Bioproject PRJNA1124409 and PRJNA951874 on the Sequence Read Archive (SRA).

## ACKNOWLEDGEMENTS

The authors would like to acknowledge Aldrin V. Imbag and Fatima Rose S. Guemo for providing and maintaining the high-performance computing resources that enabled the assembly and analysis of data, Charalyn A. Babida for managing the metadata of all samples during genomic surveillance, and Othoniel Onza for overseeing procurement processes in our laboratory. We extend our gratitude to the Department of Health Epidemiology Bureau of the Philippines, the COVID-19 Surveillance and Quick Action Unit, the Regional Epidemiology Surveillance Units, and our partner Sub-National Laboratories; Baguio General Hospital and Medical Center (BGHMC), Lung Center of the Philippines (LCP), San Lazaro Hospital (SLH), Vicente Sotto Memorial Medical Center (VSMMC), and Southern Philippines Medical Center (SPMC) for their invaluable support in the nation’s biosurveillance efforts.

## REFERENCES

Bal, A., Simon, B., Destras, G., Chalvignac, R., Semanas, Q., Oblette, A., Quéromès, G., Fanget, R., Regue, H., Morfin, F., Valette, M., Lina, B., & Josset, L. (2022). Detection and prevalence of SARS-CoV-2 co-infections during the Omicron variant circulation in France. Nature Communications, 13(1), 6316. 10.1038/s41467-022-33910-9

Bolze, A., Basler, T., White, S., Dei Rossi, A., Wyman, D., Dai, H., Roychoudhury, P., Greninger, A. L., Hayashibara, K., Beatty, M., Shah, S., Stous, S., McCrone, J. T., Kil, E., Cassens, T., Tsan, K., Nguyen, J., Ramirez, J., Carter, S.,…Luo, S. (2022). Evidence for SARS-CoV-2 Delta and Omicron co-infections and recombination. Med, 3(12), 848–859.e4. 10.1016/j.medj.2022.10.002

Combes, P., Bisseux, M., Bal, A., Marin, P., Latour, J., Archimbaud, C., Brebion, A., Chabrolles, H., Regagnon, C., Lafolie, J., Destras, G., Simon, B., Izopet, J., Josset, L., Henquell, C., & Mirand, A. (2022). Evidence of co-infections during Delta and Omicron SARS-CoV-2 variants co-circulation through prospective screening and sequencing. Clinical Microbiology and Infection, 28(11), 1503.e5-1503.e8. 10.1016/j.cmi.2022.06.030

Dezordi, F. Z., Resende, P. C., Naveca, F. G., do Nascimento, V. A., de Souza, V. C., Paixão, A. C. D., Appolinario, L., Lopes, R. S., Mendonça, A. C. F., da Rocha, A. S. B., Venas, T. M. M., Pereira, E. C., Paiva, M. H. S., Docena, C., Bezerra, M. F., Machado, L. C., Salvato, R. S., Gregianini, T. S., Martins, L. G., Pereira, F. M., Rovaris, D. B., Fernandes, S. B., Ribeiro-Rodrigues, R., Costa, T. O., Sousa, J. C., Jr., Miyajima, F., Delatorre, E., Gräf, T., Bello, G., Siqueira, M. M., & Wallau, G. L. (2022). Unusual SARS-CoV-2 intrahost diversity reveals lineage superinfection. Microbial Genomics. Retrieved from 10.1099/mgen.0.000751

Ewels, P.A., Peltzer, A., Fillinger, S. et al. (2020). The nf-core framework for community-curated bioinformatics pipelines. Nat Biotechnol 38, 276–278. Retrieved from 10.1038/s41587-020-0439-x

Hadfield, J., Bedford, T., Neher, R., Aksamentov, I., Anderson, J.S.J, Andrews, K., Chang, J., Hodcroft, E., Huddleston, J., Lee, J., Lin, V., Roemer, C., Sibley, T. (2018) Nextstrain: real-time tracking of pathogen evolution, Bioinformatics (2018). Retrieved from https://docs.nextstrain.org

Jackson, B., Boni, M.F., Bull, M.J., Colleran, A., Colquhoun, R.M., Darby, A.C., Haldenby, S., Hill, V., Lucaci, A., McCrone, J.T., Nicholls, S.M., O’Toole, Á., Pacchiarini, N., Poplawski, R., Scher, E., Todd, F., Webster, H.J., Whitehead, M., Wierzbicki, C. (2021) COVID-19 Genomics UK (COG-UK) Consortium; Loman NJ, Connor TR, Robertson DL, Pybus OG, Rambaut A. Generation and transmission of interlineage recombinants in the SARS-CoV-2 pandemic. Cell. Sep 30;184(20):5179–5188.e8. doi: 10.1016/j.cell.2021.08.014. Epub 2021 Aug 17. PMID: 34499854; PMCID: PMC8367733.

Karthikeyan, S., Levy, J.I., De Hoff, P., Humphrey, G., Birmingham, A., Jepsen, K., Farmer, S., Tubb, H.M., Valles, T., Tribelhorn, C.E., Tsai, R., Aigner, S., Sathe, S., Moshiri, N., Henson, B., Mark, A.M., Hakim, A., Baer, N.A., Barber, T., Belda-Ferre, P., Chacón, M., Cheung, W., Cresini, E.S., Eisner, E.R., Lastrella, A.L., Lawrence, E.S., Marotz, C.A., Ngo, T.T., Ostrander, T., Plascencia, A., Salido, R.A., Seaver, P., Smoot, E.W., McDonald, D., Neuhard, R.M., Scioscia, A.L., Satterlund, A.M., Simmons, E.H., Abelman, D.B., Brenner, D., Bruner, J.C., Buckley, A., Ellison, M., Gattas, J., Gonias, S.L., Hale, M., Hawkins, F., Ikeda, L., Jhaveri, H., Johnson, T., Kellen, V., Kremer, B., Matthews, G., McLawhon, R.W., Ouillet, P., Park, D., Pradenas, A., Reed, S., Riggs, L., Sanders, A., Sollenberger, B., Song, A., White, B., Winbush, T., Aceves, C.M., Anderson, C., Gangavarapu, K., Hufbauer, E., Kurzban, E., Lee, J., Matteson, N.L., Parker, E., Perkins, S.A., Ramesh, K.S., Robles-Sikisaka, R., Schwab, M.A., Spencer, E., Wohl, S., Nicholson, L., Mchardy, I.H., Dimmock, D.P., Hobbs, C.A., Bakhtar, O., Harding, A., Mendoza, A., Bolze, A., Becker, D., Cirulli, E.T., Isaksson, M., Barrett, K.M.S., Washington, N.L., Malone, J.D., Schafer, A.M., Gurfield, N., Stous, S., Fielding-Miller, R., Garfein, R.S., Gaines, T., Anderson, C., Martin, N.K., Schooley, R., Austin, B., MacCannell, D.R., Kingsmore, S.F., Lee, W., Shah, S., McDonald, E., Yu, A.T., Zeller, M., Fisch, K.M., Longhurst, C., Maysent, P., Pride, D., Khosla, P.K., Laurent, L.C., Yeo, G.W., Andersen, K.G., Knight, R. (2022) Wastewater sequencing uncovers early, cryptic SARS-CoV-2 variant transmission. Retrieved from doi: 10.1101/2021.12.21.21268143

Khare, S., et al. (2021) GISAID’s Role in Pandemic Response. China CDC Weekly, 3(49): 1049–1051. doi: 10.46234/ccdcw2021.255 PMCID: 8668406

Liao, H., Cai, D. & Sun, Y. (2022). VirStrain: a strain identification tool for RNA viruses. Genome Biol 23, 38. 10.1186/s13059-022-02609-x

Lindenbaum, P. & Redon, R. (2017). bioalcidae, samjs and vcffilterjs: object-oriented formatters and filters for bioinformatics files. Bioinformatics, Volume 34, Issue 7, April 2018, Pages 1224–1225. Retrieved from 10.1093/bioinformatics/btx734

Milne, I., Stephen, G., Bayer, M., Cock, P.J.A., Pritchard, L., Cardle, L., Shaw, P.D. and Marshall, D. (2013). Using Tablet for visual exploration of second-generation sequencing data. Briefings in Bioinformatics 14(2), 193–202. Retrieved from https://ics.hutton.ac.uk/tablet/

Molina-Mora, J. A., Cordero-Laurent, E., Calderón-Osorno, M., Chacón-Ramírez, E., & Duarte-Martínez, F. (2022). Metagenomic pipeline for identifying co-infections among distinct SARS-CoV-2 variants of concern: Study cases from Alpha to Omicron. Scientific Reports, 12(1), 9377. 10.1038/s41598-022-13113-4

O’Toole, Á., Scher, E., Underwood, A., Jackson, B., Hill, V., McCrone, J. T., Colquhoun, R., Ruis, C., Abu-Dahab, K., Taylor, B., Yeats, C., du Plessis, L., Maloney, D., Medd, N., Attwood, S. W., Aanensen, D. M., Holmes, E. C., Pybus, O. G., & Rambaut, A. (2021). Assignment of epidemiological lineages in an emerging pandemic using the pangolin tool. Virus Evolution, 7(2), veab064. 10.1093/ve/veab064

Pangilinan, E. A. R., Egana, J. M. C., Mantaring, R. J. Q., Telles, A. J. E., Tablizo, F. A., Lapid, C. M., Yangzon, M. S. L., Endozo, J. J. S., Padilla, K. S. A. R., Nipales, J. E., Carandang, L. C. D. L., Enriquez, Z. M. R., Barot, T. A. U., Manlimos, R. A., Mangonon, K. N. P., Plantig, Ma. E. L., Araiza, S. M. M., Llames, J.-H. S., Punayan, K. P.,…Saloma, C. P. (2023). Analysis of SARS-CoV-2 Recombinant Lineages XBC and XBC.1 in the Philippines and Evidence for Delta-Omicron Co-infection as a Potential Origin [Preprint]. Genetics. 10.1101/2023.04.12.534029

Pedro, N., Silva, C.N., Magalhães, A.C., Cavadas, B., Rocha, A.M., Moreira, A.C., Gomes, M.S., Silva, D., Sobrinho-Simões, J., Ramos, A., et al. (2021) Dynamics of a Dual SARS-CoV-2 Lineage Co-Infection on a Prolonged Viral Shedding COVID-19 Case: Insights into Clinical Severity and Disease Duration. Microorganisms. 9(2):300. 10.3390/microorganisms9020300

Perez-Florido, J., Casimiro-Soriguer, C. S., Ortuño, F., Fernandez-Rueda, J. L., Aguado, A., Lara, M., Riazzo, C., Rodriguez-Iglesias, M. A., Camacho-Martinez, P., Merino-Diaz, L., Pupo-Ledo, I., de Salazar, A., Viñuela, L., Fuentes, A., Chueca, N., The Andalusian COVID-19 Sequencing Initiative, García, F., Dopazo, J., & Lepe, J. A. (2023). Detection of High Level of Co-Infection and the Emergence of Novel SARS CoV-2 Delta-Omicron and Omicron-Omicron Recombinants in the Epidemiological Surveillance of Andalusia. International Journal of Molecular Sciences, 24(3), 2419. 10.3390/ijms24032419

Pipek, O.A., Medgyes-Horváth, A., Stéger, J. et al. (2024). Systematic detection of co-infection and intra-host recombination in more than 2 million global SARS-CoV-2 samples. Nat Commun 15, 517. Retrieved from 10.1038/s41467-023-43391-z

Pisano, M.B., Sicilia, P., Zeballos, M., Lucca, A., Fernandez, F., Castro, G.M., Goya, S., Viegas, M., López, L., Barbás, M.G. and Ré, V.E. (2022) SARS-CoV-2 Genomic Surveillance Enables the Identification of Delta/Omicron Co-Infections in Argentina. Front.Virol. 2:910839. doi: 10.3389/fviro.2022.910839

Quick, J. (2020).nCoV-2019 sequencing protocol v3 (LoCost) V.3. Retrieved from 10.17504/protocols.io.bp2l6n26rgqe/v3

Rockett, R. J., Draper, J., Gall, M., Sim, E. M., Arnott, A., Agius, J. E., Johnson-Mackinnon, J., Fong, W., Martinez, E., Drew, A. P., Lee, C., Ngo, C., Ramsperger, M., Ginn, A. N., Wang, Q., Fennell, M., Ko, D., Hueston, L., Kairaitis, L.,…Sintchenko, V. (2022). Co-infection with SARS-CoV-2 Omicron and Delta variants revealed by genomic surveillance. Nature Communications, 13(1), 2745. 10.1038/s41467-022-30518-x

Ruis, C. (2022). bammix: Summarise nucleotide counts at a set of positions in a BAM file to search for mixtures. Retrieved from https://github.com/chrisruis/bammix

Samoilov, A.E., Kaptelova, V.V., Bukharina, A.Y. et al. (2021). Case report: change of dominant strain during dual SARS-CoV-2 infection. BMC Infect Dis 21, 959 Retrieved from 10.1186/s12879-021-06664-w

Schimmel, L., Rohwer, K., Poon, A., & Symalla, T. (2022). Sc2rf - SARS-CoV-2 recombinant finder. Retrieved from https://github.com/lenaschimmel/sc2rf

Tamura, T., Ito, J., Uriu, K. et al. (2023). Virological characteristics of the SARS-CoV-2 XBB variant derived from recombination of two Omicron subvariants. Nat Commun 14, 2800. Retrieved from 10.1038/s41467-023-38435-3

Tonkin-Hill, G., Martincorena, I., Amato, R., Lawson, A. R. J., Gerstung, M., Johnston, I., Jackson, D. K., Park, N., Lensing, S. V., Quail, M. A., Gonçalves, S., Ariani, C., Chapman, M. S., Hamilton, W. L., Meredith, L. W., Hall, G., Jahun, A. S., Chaudhry, Y., Hosmillo, M., Pinckert, M. L., Georgana, I., Yakovleva, A., Caller, L. G., Caddy, S. L., Feltwell, T., Khokhar, F. A., Houldcroft, C. J., Curran, M. D., Parmar, S., The COVID-19 Genomics UK (COG-UK) Consortium, Alderton, A., Nelson, R., Harrison, E. M., Sillitoe, J., Bentley, S. D., Barrett, J. C., Torok, M. E., Goodfellow, I. G., Langford, C., Kwiatkowski, D., & the Wellcome Sanger Institute COVID-19 Surveillance Team. (2021). Patterns of within-host genetic diversity in SARS-CoV-2. eLife, 10, e66857. Retrieved from 10.7554/eLife.66857

Trémeaux, P., Latour, J., Ranger, N., Ferrer, V., Harter, A., Carcenac, R., Boyer, P., Demmou, S., Nicot, F., Raymond, S., & Izopet, J. (2023). SARS-CoV-2 co-infections and recombinations identified by long-read single-molecule real-time sequencing. Microbiology Spectrum, 11(e00493-23). 10.1128/spectrum.00493-23

Turakhia, Y., Thornlow, B., Hinrichs, A. et al. (2022). Pandemic-scale phylogenomics reveals the SARS-CoV-2 recombination landscape. Nature 609, 994–997. 10.1038/s41586-022-05189-9

World Health Organization (2024). Initial risk evaluation of XEC, 09 December 2024. Retrieved from https://www.who.int/docs/default-source/coronaviruse/09122024_xec_ire.pdf

Zhou, H.-Y., Cheng, Y.-X., Xu, L., Li, J.-Y., Tao, C.-Y., Ji, C.-Y., Han, N., Yang, R., Li, Y., & Wu, A. (2021). Genomic evidence for divergent co-infections of SARS-CoV-2 lineages. Microbiology. 10.1101/2021.09.03.458951

